# Exploring the implementation and integration of structured medication reviews in primary care: A qualitative evaluation using normalization process theory

**DOI:** 10.1101/2025.08.21.25334150

**Authors:** Claire Reidy, Anna Seeley, Katherine Tucker, Paul A Bateman, Christopher E Clark, Andrew Clegg, Gary A Ford, Seema Gadhia, William Hinton, F D Richard Hobbs, Sundus Jawad, Kamlesh Khunti, Gregory Y H Lip, Simon de Lusignan, Jonathan Mant, Deborah McCahon, Bernardo Meza-Torres, Rupert A Payne, Rafael Perera-Salazar, Samuel Seidu, James P Sheppard, Marney Williams, Cynthia Wright Drakesmith, Richard J McManus, Rebecca K. Barnes

**Affiliations:** Nuffield Department of Primary Care Health Sciences, University of Oxford, Oxford, UK; Exeter Collaboration for Academic Primary Care, University of Exeter, Exeter, UK; Academic Unit for Ageing & Stroke Research, University of Leeds, Leeds, UK; Bradford Institute for Health Research, Bradford Teaching Hospitals NHS Foundation Trust, Bradford, UK; Radcliffe Department of Medicine, University of Oxford, Oxford, UK; Oxford University Hospitals NHS Foundation Trust, UK; Health Innovation Oxford and Thames Valley, UK; NHS Frimley Integrated Care Board, UK; Diabetes Research Centre, University of Leicester, Leicester, UK; Liverpool Centre for Cardiovascular Science at University of Liverpool, UK; Liverpool John Moores University and Liverpool Heart & Chest Hospital, Liverpool, UK; Danish Center for Health Services Research, Department of Clinical Medicine, Aalborg University, Aalborg, Denmark; Department of Cardiology, Lipidology and Internal Medicine with Intensive Coronary Care Unit, Medical University of Bialystok, Bialystok, Poland; Primary Care Unit, Department of Public Health & Primary Care, University of Cambridge; Centre for Academic Primary Care (CAPC), Population Health Sciences, Bristol Medical School, University of Bristol; Patient and Public Involvement Representative, Nuffield Department of Primary Care Health Sciences, University of Oxford, Oxford, UK; Brighton and Sussex Medical School, University of Brighton and University of Sussex, Brighton

**Keywords:** General Practice, Clinical Pharmacists, Structured Medication Reviews, Medicines Optimisation, Normalization Process Theory, Polypharmacy

## Abstract

**Background:** Structured Medication Reviews (SMRs) were introduced into primary care in England for patients living with multiple long-term conditions (MLTCs), polypharmacy, increased frailty, in care homes or at risk of medicines-related harm. SMRs aim to optimise the therapeutic potential of medication and reduce medicine-related harms through holistic reviews.

**Aim:** To explore the day-to-day work being undertaken with, and by, clinical pharmacists to implement, embed and integrate SMRs into practice, and consider how to optimise SMRs.

**Design and setting:** Qualitative one-to-one interviews with clinical pharmacists undertaking SMRs and SMR service leaders/managers (SMR leads) in England between February 2023 and November 2024.

**Method:** Participants were recruited as part of a wider evaluation of the roll-out of SMRs in England. Interview topic guides and qualitative data analysis were informed by Normalization Process Theory (NPT).

**Results:** Eighteen clinical pharmacists and five SMR leads participated. Participants reported often having to explain the purpose of SMRs and clinical pharmacists’ roles to patients, partly due to patients not being informed about SMRs. Participants valued SMRs and expressed that trust-building and tailored consultations were important for optimising medications. Integration varied due to high workload, inconsistent leadership support, inadequate administrative/pharmacist technician resource and lack of training. However, participants described SMRs as valuable for identifying and addressing unmet needs and supporting holistic, person-centred care across MLTC pathways.

**Conclusion:** The findings demonstrate the need for improved information on SMRs for patients and primary care teams, adequate and appropriate resource allocation, and enhanced support for consultation skills training to optimise medicines use.

**How this fits in:**

- SMRs were formally introduced to primary care in 2020 to address the challenges of managing polypharmacy in an ageing population with increasing patient complexity and MLTCs.
- SMRs were introduced alongside the expansion of clinical pharmacist roles in General Practice as a comprehensive, person-centred review of all a patient’s medicines.
- This qualitative evaluation examines the day-to-day work of implementing and embedding SMRs, highlighting challenges to implementation and integration.
- Our findings reveal challenges to the sustainability of SMRs and identify opportunities for optimisation, including addressing pharmacists’ training needs and resource allocation for administrative and pharmacy technician support.

## Introduction

Medicines-related harm poses a significant burden on patients and the NHS. Adverse drug events (ADEs), which include medication errors, adverse drug reactions, allergic reactions, and overdoses, are estimated to cost the NHS approximately £400 million annually in hospital admissions alone [1]. Of the estimated 237 million medication errors occurring annually in England, nearly 40% take place in primary care and over 40% in care homes [2]. Although prescribed medications play a vital role in improving health outcomes, and prescribing multiple medications (“polypharmacy”[3]) is often clinically necessary, older adults are more likely to be prescribed multiple medications [4] and are more susceptible to harm from medicines [5]. Such harm includes hospitalisation, falls, drug interactions, and mortality, especially for those living with frailty or multiple long-term conditions (MLTCs) [4-9]. Structured Medication Reviews (SMRs) have been introduced to help manage polypharmacy, particularly for patients with MLTCs [10, 11].

During 2020/2021, SMRs became a contractual requirement under the NHS England Network Contract Directed Enhanced Service (DES) for the then newly created Primary Care Networks (PCNs) in England [12]. The DES contract included delivering SMRs through appropriately trained healthcare professionals (primarily, clinical pharmacists) [13]. This aligned with the expansion of this role through the Additional Roles Reimbursement Scheme and the Impact and Investment Fund (IIF), a financial incentive scheme rewarding performance against national prescribing and prevention targets [12].

SMRs were targeted at patients with severe frailty, in care homes, taking 10 or more medications, medicines associated with medication errors or addictive medications [13]. They offer a more comprehensive, structured, and holistic approach to medicines management [14], as opposed to prioritising efficiency over thoroughness [15]. It was hoped that by providing individual and holistic reviews of medications, pharmacists could optimise medication use, aligned with patients’ needs and preferences [13, 16-20].

This evaluation explores the experiences of clinical pharmacists and SMR service leaders and managers a few years on from SMR roll-out. Informed by Normalization Process Theory (NPT) [21], we focus on the everyday work undertaken by organisations and individuals to implement, embed and integrate SMRs into general practice at a PCN and practice level, and to consider how SMRs might be optimised and sustained.

## Methods

### Study design

Semi-structured qualitative interviews were undertaken as part of a wider mixed-methods evaluation (Optimising Structured Medication Reviews - OSCAR) to explore the implementation and roll-out of SMRs in England.

### Participant recruitment and sampling

We spoke to clinical pharmacists undertaking SMRs in primary care settings, and SMR service/medicines optimisation leaders and managers (referred to as “SMR leads”), working within different settings. Some participants held dual clinical and leadership roles. For participant recruitment, those who left their details in a survey of SMRs as part of the wider OSCAR evaluation [22] were contacted, while others working in OSCAR case study sites were invited to participate by the research team via email. All participants gave written or digitally recorded informed consent.

### Data collection

All interviews were conducted by AA (PhD, post-doctoral researcher), AS (GP and primary care DPhil student) and RB (PhD, experienced qualitative researcher). Neither AA, nor RB, had any prior relationship with the study participants. AS had previously worked briefly with one of the participants but was not working with them at the time of interview. Interviews took place on Microsoft Teams or via telephone between February 2023 and November 2024.

Flexible topic guides (see Supplementary documents A and B), sensitised to the constructs of NPT, were developed with input from clinicians and patients. Interviewees were asked to reflect on the different kinds of work required in preparing for, conducting, and implementing SMRs, including resources used and how different patient groups were prioritised. Data collection continued until no new insights regarding the research questions and theoretical framework were presented [23]. All interviews were digitally recorded and professionally transcribed verbatim.

### Data analysis

#### Theoretical framework

NPT is a methodological framework that aids in exploring the implementation of healthcare practices as well as providing feedback for helping to sustain further implementation [21]. In the context of SMRs, NPT considers the implementation mechanisms and the collaborative work involved in integrating SMRs into routine healthcare practice, in terms of personal and group resources [24]. NPT can shed light on the unseen work involved in embedding SMRs within a healthcare system already challenged by competing demands [25]. The four key NPT constructs are outlined in Table 1.

**Table 1.** NPT Constructs.

| NPT Constructs | Interpretation |
| --- | --- |
| Coherence | Addresses the question, “What is the work?”. It involves the sense-making processes that individuals and teams undertake to understand and define the purpose and tasks needed to integrate SMRs into routine practice. |
| Cognitive participation | Asks “Who does the work?” and explores how individuals and organisations engage with and commit to a new practice and considers buy-in and ongoing relational work. |
| Collective action | Focuses on “How does the work get done?” and highlights the operational activities needed to implement SMRs, and how these practices are integrated into workflows. |
| Reflexive monitoring | Is essential for embedding and maintaining clinical practices. It represents how individuals and teams evaluate the impact and value of SMRs, and the continued refinement of SMRs. |

Following transcription, interview records were anonymised and uploaded to NVivo (v14). The data were coded both inductively, guided by Braun and Clarke’s [26] five-step framework for thematic analysis, and deductively, using the primary NPT constructs and secondary constructs [25]. Firstly, CR (experienced primary healthcare researcher and with experience of NPT) became familiar with the data through multiple readings of a subset of transcripts, creating an initial list of ideas from the data. These steps created initial codes, which were then reviewed together with RB. This was followed by inductive and deductive development of broad themes within the NPT constructs. Emerging findings were presented to the wider study team for sense-checking and discussion. The wider study team comprised multiple members with varying clinical and research backgrounds, and patient and public involvement, enabling investigator triangulation [27].

## Results

Interviews were conducted with 23 participants, 18 of whom were clinical pharmacists (or senior clinical pharmacists) and 5 were “SMR leads” (see Table 2). Participants were from different 7 NHS England regions and interviews lasted between 26 and 65 minutes (average□=□37 minutes).

**Table 2:** Participant characteristics (n=23)

|  |  |  |
| --- | --- | --- |
| Age N (%) | 20-29 | 3 (13%) |
|  | 30-39 | 11 (48%) |
|  | 40-49 | 7 (30%) |
|  | 50-59 | 2 (9%) |
| Gender | Female | 15 (65%) |
|  | Male | 8 (35%) |
| Ethnicity | British South Asian | 11 (48%) |
|  | White British | 7 (30%) |
|  | Black African | 2 (9%) |
|  | Vietnamese/Chinese | 1 (4%) |
|  | Mixed British | 1 (4%) |
|  | White Other | 1 (4%) |
| Location | North West | 5 (22%) |
|  | East Midlands | 4 (17%) |
|  | West Midlands | 2 (9%) |
|  | London | 2 (9%) |
|  | South Central | 5 (22%) |
|  | South East | 3 (13%) |
|  | South West | 2 (9%) |
| Role | PCN-based Clinical Pharmacist | 10 (44%) |
|  | GP Practice-based Clinical Pharmacist | 4 (17%) |
|  | Senior or Lead Pharmacist | 4 (17%) |
|  | PCN Pharmacy Lead | 2 (9%) |
|  | Social and Research Lead GP Partner | 1 (4%) |
|  | Care Homes Medicines optimisation Lead | 1 (4%) |
|  | Pharmacy Lead | 1 (4%) |
| Employer | PCN | 11 (48%) |
|  | GP Practice | 7 (30%) |
|  | Community Interest Company | 2 (9%) |
|  | NHS England and PCN | 1 (4%) |
|  | NHS England and ICB | 1 (4%) |
|  | ICB | 1 (4%) |
| Independent prescriber (IP) | Yes | 19 (83%) |
|  | No | 3 (13%) |
|  | GP | 1 (4%) |
| Years qualified as a pharmacist (Mean (SD*), range) | 13.65 years (SD=7.2), 2-26 years |  |
| Years experience of conducting SMRs (Mean (SD), range) | 3.9 years (SD=3.7), 0.5 – 20 years** |  |

\*Standard deviation \*\*SMRs were put into practice in 2020 but some Pharmacists described doing what they considered to be SMRs before this time.

Informed by NPT, the themes identified are summarised in Table 3, and illustrative quotes are provided.

**Table 3:**
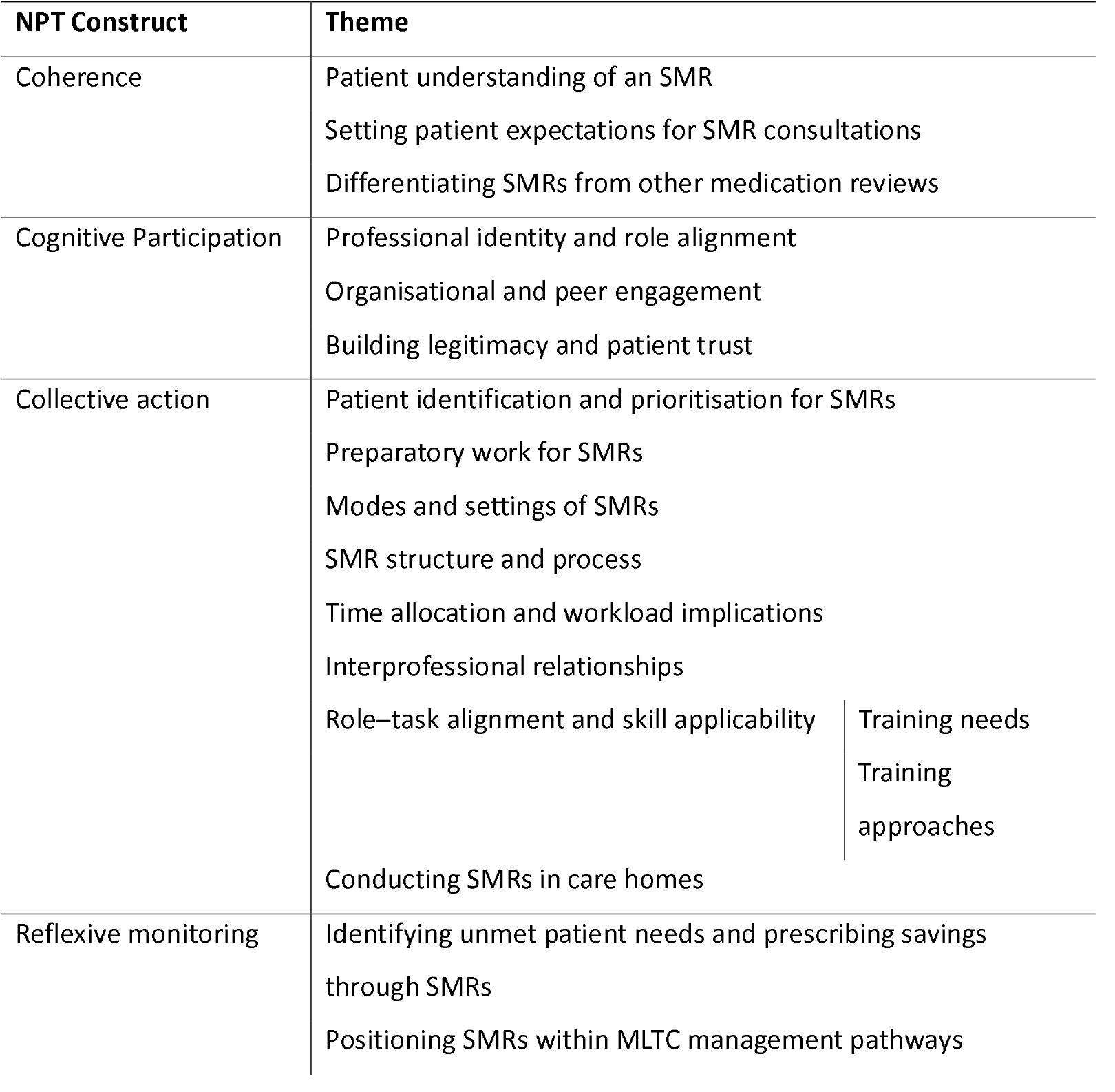
Themes by NPT constructs.

### Coherence

#### Patient understanding of an SMR

Participants often highlighted the necessity of explaining the role and purpose of an SMR to patients. They remarked that many patients still had difficulty differentiating between community pharmacists and clinical pharmacists working in primary care. This meant that clinical pharmacists often had to explain their role and how it differed:

> *“And most of them will say, “I didn’t know they had a pharmacist attached to the practice.” And then often you have to say to them, “No, I’m not from your local pharmacy,” because once you say pharmacy they think you’re from the community pharmacy.” (PHR 10, qualified=19 years, SMRs=6 years, IP)*

It was reported that, potentially due to a lack of understanding, some patients viewed the SMR as a procedural exercise to continue access to repeat medications. Some interviewees supposed that providing patients with more information about SMRs beforehand would improve the value attributed to SMRs.

#### Setting patient expectations for SMR consultations

Some practices offered patients booked appointments for SMRs in advance while others did not, which participants described as disconcerting for patients. Most of our participants wanted to give patients information and time to prepare, which could reduce time spent explaining the purpose of SMRs and allow greater focus on medication needs and queries:

> *“you do really need to get somebody an appointment because I want them to anticipate it so they can think about any questions as well…cold calling…I don’t think that’s the best approach.” (PHR 16, qualified=24 years, SMRs= 6 years, IP)*

#### Differentiating SMRs from other medication reviews

Many of the pharmacists spoke of SMRs as holistic reviews, which provide the potential to prescribe better for patients. They referred to SMRs as more complex and thorough than GP-led medication reviews, yet expressed that other clinicians sometimes lacked this comprehension:

> *“I think it’s a skill thing, you ask a pharmacist what a SMR is and you’ll get a completely different story to what a GP says… So some GPs will see it as a tick box. “I’ve had a look through the medicines and yeah… I’ve done a SMR”… [but] you didn’t actually speak to them.” (PHR 05, qualified=26 years, SMRs=4 years, IP)*

### Cognitive Participation

#### Professional identity and role alignment

Those interviewed were confident in the role of clinical pharmacists to appropriately and thoroughly review multiple medications in-depth across several conditions. Some described how they were able to pick up on different and additional details to GPs, and many had extensive experience in relevant clinical specialities e.g. frailty:

> *“But yeah, I mean, it, it’s stuff that we, we do all the time, isn’t it. As pharmacists we talk about medicines. It’s sort of our bread and butter, really.” (PHR 01, qualified=7 years, SMRs=3.5 years, IP)*

Interviewees also spoke of holistic SMRs as useful touchpoints for pharmacists to engage with and manage MLTCs:

> *“you’re saying, asthma is this part… come back next week and we’ll do your diabetes… it feels like we’re chopping a person up into their little conditions. Whereas… their health is a holistic thing.” (PHR 01, qualified=7 years, SMRs=3.5 years, IP)*

#### Organisational and peer engagement

Some interviewees described a lack of senior buy-in, resulting in inadequate resources for SMRs (e.g., costing in pharmacist technicians, administrative support or enough pharmacist time), making it difficult to see most patients who could benefit from one. One SMR lead spoke of working with the PCN Clinical Director and PCN Board to negotiate pharmacist time allocated to SMRs:

> *“We looked…[at] how many people could be eligible for an SMR in line with what the contract says…45,000 people… across three practices…the challenge is where you start to really stratify that…also what percentage of the, the whole time equivalent that pharmacists have…versus every other thing they do.” (LDR 03, qualified=12 years, SMRs=3 years)*

Where adequate resources had been allocated, interviewees reported being able to appropriately tailor their week (e.g., holding weekly SMR clinics, booking patients in, arranging/checking blood tests). They could then focus their time better.

Many participants suggested that a lack of engagement and involvement from other clinicians was perhaps due to the lack of understanding of the purpose and value of SMRs. However, more established pharmacists described building good rapport with colleagues in their practice and improving appreciation of the length and breadth of SMRs, over time. This was especially relevant for PCN-based pharmacists who covered several practices.

#### Building legitimacy and patient trust

Those interviewed described an apparent expectation from patients that GPs held the primary role in discussing medications. There was a resulting aspiration from interviewees to build legitimacy and trust with patients:

> *“sometimes when they [patients] hear…that you’re a pharmacist and trying to make changes to their medication they sometimes aren’t really too susceptible to it because they are, “why isn’t the GP doing it and why hasn’t the GP noticed that before?” (PHR 07, qualified=2 years, SMRs=2 years)*

Many proposed that patients’ conviction in the pharmacist role and purpose of SMRs impacted how open patients were in discussing their medications and whether they felt able or willing to consider changes to their existing regimen.

### Collective action

#### Patient identification and prioritisation for SMRs

Interviewees described undertaking SMRs with patients from the priority cohorts set out in the DES contract, and some also targeted patients on specific medication combinations or with medications with potentially harmful omissions outlined in the 2022/23 IIF. Many of these patients were identified via Ardens, Eclipse or EMIS searches (proactive, automated searches of the practice’s electronic health records), with others identified opportunistically. These systems worked well when there were clear criteria for patient prioritisation however, some practices ceased to prioritise SMRs after the withdrawal of IIF incentives to undertake SMRs. Alternatively, some interviewees depicted practices or PCNs as acting more autonomously, focusing on local priorities for SMRs.

#### Preparatory work for SMRs

Interviewees described undertaking extensive preparatory work to “build a picture” before speaking to patients. This involved determining what condition(s) the patient had and understanding what was going on in their lives medically (through hospital letters, reported medication changes, blood tests, including liver function, cholesterol tests, and blood pressure readings). Also, assessment of medication prescribing patterns (e.g., what is taken, and when requested, if at all), and inappropriate combinations. Preparatory work enabled better understanding of the complexity and history of the patient:

> *“I’ve made myself a little checklist so, I do a lot of work pre-calling in the patient…I look for like, you know, the blood tests have like, for example, if it is a cholesterol medication, I check whether their liver function has been done…so then I can just build a picture before I actually speak to them.” (PHR 08, qualified=11 years, SMRs=3 years)*

This preparation could take 10-20 minutes and aimed to optimise and focus SMR time. It was also noted that this work reduced over time as pharmacists knew the patients better (so needed less preparatory work) and patients were more prepared.

Importantly, however, blood test results were often not undertaken before an SMR, which made initiating changes during an SMR difficult or impossible, and so follow-up appointments were required to initiate, or even consider, some changes.

#### Modes and settings of SMRs

SMRs were undertaken in the practice, care homes or patients’ homes or remotely. The majority were conducted remotely by telephone due to time efficiencies or lack of clinic space. Face-to-face SMRs were preferred for some older or more complex patients (especially to assess frailty), or for patients with English language difficulties, hearing difficulties, or when others (e.g., carers/family members) were involved. Face-to-face SMRs were described as better at determining whether patients understand their medications, for example by having access to facial expressions and other non-verbal cues:

> *“I would prefer a face-to-face SMR where patients come in, bring in all their medication with them*..*[and] understanding of their body language, [and] whether they take their medication appropriately. (PHR 03, qualified=30 years, SMRs=3 years, IP)*

#### SMR structure and process

After establishing the purpose of the appointment, interviewees described going through patients’ test results (if available) and medications one by one, often while following a clinical consultation guide/”template” (e.g., a multimorbidity, self-made or generic electronic template). Templates were used as a prompt and to initiate broader, open questions, e.g., to understand how medication is taken and any side effects experienced. Interviewees reported exploring social circumstances and questions or concerns about the medication or other health matters. Then, coming to a shared agreement about any changes (e.g. medication doses, adding or removing medication). Where changes were agreed, these sometimes resulted in further follow-up (especially if dependent on blood tests):

> *“I have my own set of questions…have you got a good routine for your medication? Any over the counter medication…we normally go through all each and every single medication. We go through their concerns first…[and] try to come to sort of shared agreement with regards to a plan…that can result in another follow-up appointment.” (PHR 07, qualified=2 years, SMRs=2 years)*

While existing templates were useful to aid the process of SMRS, they were also seen as very detailed, difficult to navigate around and time-consuming to follow, especially for those newer to the role:

> *“I’ve got used to the template now. So, I, I find it works quite well for me. I think it’s a case of getting used to it. When I first started, I found it quite hard to navigate around it. You make it work right, because that’s the only template you have.” (PHR 10, qualified=19 years)*

#### Time allocation and workload implications

SMRs were typically described as lasting at least 20 minutes, and often extended to 30 or 45 minutes or more, depending on the complexity of patients’ needs. Some also required follow-up consultation(s). The time allocated for SMRs, compared to standard GP appointments, was seen as crucial in building rapport and trust, and allowing deeper exploration of patient complexity. Interviewees said this time enabled patients to reflect on their medication use and to speak openly, providing more opportunity for partnership and shared decision-making:

> *“there was a lot of to-ing and fro-ing about how long we were gonna be allowed to do them in…so I stood my ground and still kept half an hour for an SMR plus twenty minutes follow-up [if needed]… in some areas pharmacists have ended up doing them in a lot shorter time, that doesn’t give the patient as much freedom I think to actually be honest with you and kind of get that rapport built.” (LDR 04, qualified=14 years, SMRs=3 years)*

Giving patients sufficient time to express their concerns was frequently described as essential to conducting an SMR well, ensuring that patients’ perspectives were being heard and addressed.

#### Interprofessional relationships

Where interviewees did not have the authority or confidence to change an existing medication or start a new one (e.g., not an independent prescriber (IP) or unfamiliar with a particular medication/condition), it was resolved with a GP. This relied on collaborative relationships with GPs in the practice:

> *“if the SMR is okay…then I wouldn’t, wouldn’t share that with the GP. It tends to be now more if there are sort of medication changes or new medicines that need to be prescribed, that’s when…[to] involve or …with regards to symptoms…I would sort of have a chat with the GP or debrief.” (PHR 04, qualified=14 years, SMRs=2 years)*

#### Role–task alignment and skill applicability

##### Training needs

Interviewees appeared to be familiar and comfortable with making changes with medications that matched their own professional knowledge and experience. However, some changes were reported as being difficult, largely concerning pain and addictive medications. Any dose changes here were depicted as a slow and difficult process, requiring careful conversations, advanced consultation skills, and multiple follow-ups:

> *“it’s resistance to people wanting to bring themselves off pain relief medication and… I mean refusing tablet doses to be reduced. So, despite being in their best interests, they see it differently…so it’s a question of understanding and educating them and that can be potentially problematic as well.” (PHR 06, qualified=10 years, SMRs=3.5 years, IP)*

While pharmacists expressed that conducting SMRs fitted their role, some training needs were identified to prepare newer clinical pharmacists and to optimise SMR delivery. For example, expertise to guide and support patients living with particular conditions (such as Parkinson’s Disease), on particular medications or engaging in difficult conversations (especially around deprescribing).

Interviewees commonly highlighted training needs, often centred around building confidence and consultation skills for stopping or reducing certain medications. These were particularly around building rapport and trust with patients, encouraging open conversations, and supporting patient disclosure. Additional priorities included shared decision-making, interpreting non-verbal cues, and developing knowledge and confidence in deprescribing for patients in care homes due to deteriorating physiology and increased frailty. Mental health skills and consultation length and structure were also noted.

#### Training approaches

Interviewees described the training they had undertaken to prepare them for conducting SMRs (see Box 1). There was a huge variation in the types of training described (which largely depended on the PCN or practice) and also the time allocated to receive training.

##### Box 1.

*Training approaches described by interviewees*

###### Shadowing

Shadowing a more senior and experienced clinician gave insight into how consultations were conducted, providing a model of how to carry them out, especially for more complex cases, how templates are used, and use of clinical systems. They provided examples of more straightforward SMRs, leading up to more complex ones.

###### Leaders providing monthly support and training sessions

One PCN leader described giving support once a month to clinical pharmacists undertaking SMRs for the first time. They gave them opportunities to practice speaking with patients about medicines and sitting with them while they undertook SMRs. They also encouraged clinical pharmacists to come back and ask further questions, building their skills gradually, to support more complex patients.

###### Specific SMR training - Centre for Pharmacy Postgraduate Education pathway

One interviewee mentioned the Centre for Pharmacy Postgraduate Education (CPPE) pathway which covered their first 18 months in primary care, with training in medical conditions and SMR consultations, through workshops and case studies.

###### Creating communities of practice

One PCN lead spoke of the creation of a community of practice of those with a shared interest, from different disciplines (e.g., GPs, clinical pharmacists, practice nurses, commissioners) in “addressing and tackling problematic polypharmacy”, and where opioids are discussed.

###### Mentoring and supervision

- Some spoke of working with others to train and build confidence to undertake SMRs, and watching others do SMRs, especially with potentially more difficult scenarios, such as supporting patients to reduce and stop opioids
- One PCN provided a mentorship scheme, with training and webinars to support clinical pharmacists undertaking SMRs providing a place to answer queries, build expertise, confidence and knowledge

###### SMR training day

With the aim of growing clinical knowledge, as well as the confidence of GPs in the PCN

###### Workshops

Workshops delivered nationally, e.g. on managing opioid medications

### Conducting SMRs in care homes

Care home SMRs were widely perceived as resource-intensive. The complexity of patients, many of whom have significant cognitive impairment, along with the need to engage multiple parties (including patients, families, carers, specialist staff), was considered challenging:

> *“I think with, for care home patients, it’s the time that’s needed and that’s because there, it’s so, there’s so many people involved in their care.” (LDR 05, qualified=23 years, SMRs=20 years, IP)*

There was considerable variability in the management of SMRs in care homes. In some areas, GPs were designated to manage care home residents due to the complexities of these patients. Some PCNs assigned pharmacists to specific care homes, designated particular care homes to individual practices, or a single practice took responsibility for all care homes within the PCN.

One barrier was limited WiFi in care homes, which obstructed access to patient records. To facilitate the administration of SMRs, some care homes allocated tasks (e.g. logging patients’ blood pressure/weight) to night staff, to copy and paste into relevant records. Medication changes were often reported as well-managed within care home protocols, particularly when aligned with their monthly medication cycles. When pharmacists engaged with and aligned with these procedures and protocols, and were familiar with care home staff, SMRs ran more smoothly.

### Reflexive monitoring

#### Identifying unmet patient needs and prescribing savings through SMRs

Two SMR leads described how they carried out local service evaluations or audits of their SMRs, with evidence that SMRs facilitated prescribing savings for the practice, the identification of work to help patients and prevented hospital admissions:

> *“I’ve done a bit of a like a service evaluation… it’s led to some savings, but generally it’s led to some kind of intervention… It identifies work that we could, we needed to do…quite a few of the GPs…have said*… *[it’s] what they were hoping for, not that they would do less, but the things that they were missing would get picked up.” (LDR 02, qualified=19 years, SMRs=3 years, IP)*
>
> *“When we audited our own SMRs…we were showing a [prescribing] savings of around £280 [per year] per patient…We showed that actually we prevented a number of hospital admissions as well*…*if you found the same problem in ten people…every ten you’ve prevented one hospital admission…[but] we are quite an experienced team.” (LDR 05, qualified=23 years, SMRs=20 years, IP)*

Interviewees described how SMRs helped identify overlooked/misattributed clinical issues, review long-neglected medications, optimise symptom management, and reduce medication burden and drug interactions. Establishing trust, continuity and holistic care enabled them to support patients, such as those with complex pain, through safe, gradual medication reductions.

### Positioning SMRs within MLTC management pathways

Interviewees saw SMRs as an important element of MLTC management, providing a holistic and patient-centred approach to reviewing medications while addressing broader aspects of care. SMRs allowed assessment of advice previously provided and clarifying whether patients fully understood medication guidance, while agreeing on strategies moving forward. SMRs also provided an opportunity to consolidate primary and secondary care medication management plans, supporting patients to feel more informed and have clearer understanding of their medications:

> *“We’ve just incorporated [SMRS] as part of our…Chronic Disease Process because that is the backbone as a pharmacy team of what we do…I think it just comes down to SMRs are really important. They are really good holistic reviews and they just support a really good prescribing process.” (PHR 13, qualified=11 years, SMRs=3 years, IP)*

## Discussion

### Summary of findings

In exploring the mechanisms involved in implementing and embedding SMRs into routine primary care, interviewees reported that patients’ understanding of the purpose of SMRs and the clinical pharmacist’s role was often limited. A lack of communication or preparation and ‘cold calling’ appointments posed challenges to meaningful patient engagement.

Pharmacists demonstrated strong professional identity and confidence in delivering SMRs, particularly when supported by sufficient resources and organisational buy-in (e.g., allocation of pharmacy technician support for blood test booking and preparation). Building trust with patients and clarifying the clinical pharmacist’s role were crucial for fostering engagement, as was relationship-building with colleagues, especially for PCN-based pharmacists working across multiple practices.

Successful implementation involved complex operational work, including pre-SMR preparation and follow-ups. SMRs required adequate time, clinical knowledge, and skilled communication, particularly when deprescribing, complex cases and with care home residents. However, variability in workflows, training and resource allocation meant inconsistent integration across practices. Ongoing professional development and protected time supported safer prescribing, shared decision-making and more sustainable implementation.

Service leaders and managers used local audits and evaluations to assess impact, reporting benefits such as uncovering unmet patient needs, timely interventions, prescribing cost savings, and reductions in hospital admissions. SMRs were widely viewed as valuable opportunities to engage with patients, clarify care plans, improve patient understanding and support the management of MLTCs.

### Strengths and limitations

Using NPT enabled insights into the administrative, clinical and relational work pharmacists undertake in implementing and conducting SMRs. It also illuminated how this work is shaped by evolving policy, resource constraints and competing priorities. The study included a diverse sample of participants, varying in age, sex, ethnicity (both closely aligning with national figures [28]), role, experience, and geography. This heterogeneity enhanced the breadth of perspectives captured. However, most interviewees were IPs with several years of experience, which may limit the transferability of findings to less experienced pharmacists.

Despite this, there was strong consensus around training needs, particularly regarding consultation skills. Given the collaborative nature of SMR delivery, perspectives from GPs, practice managers, pharmacy technicians, and administrative staff could have further enriched understanding of interprofessional dynamics, buy-in, relational and operational challenges. Trustworthiness and credibility of the findings were enhanced through investigator triangulation, regular team discussions, and involving stakeholders in study design and interpretation. Finally, the findings reflect a more mature phase of roll-out and a national programme that continues to evolve in a constantly shifting policy and operational context, and so continued evaluation as the landscape changes is recommended.

### Comparison with existing literature

Our findings reflect broader evidence on workforce planning in primary care, particularly the importance of structural and organisational support in embedding new clinical roles [29, 30]. Although the integration of clinical pharmacists has been widely studied [29-32], persistent challenges remain, most notably around role clarity, team collaboration, funding security, and demonstrating economic value [31]. IP status is particularly valued by patients, and GPs are more likely to be supportive when they have experience working alongside pharmacists [32]. Building trust with both colleagues, patients and carers is therefore crucial, as is access to appropriate training, sustained funding, and clear models of integration [30, 32, 33].

In the context of SMRs, existing literature has shown a similarly mixed picture. Roll-out has been inconsistent, with pharmacists already embedded in practice adapting more easily, and newer pharmacists often relying on templates and established habits [34]. These findings echo our own, especially around the tensions pharmacists face in collaborative working, time constraints and balancing guidelines with individual patient needs, particularly in deprescribing [34, 35]. Managing medication changes in polypharmacy is often a process of ongoing adjustments and relational work, rather than one-off interventions [35], and requires preparatory and follow-up work.

Training is a recurring theme, with previous studies noting that SMR training tends to emphasise clinical knowledge but offers limited feedback and support on consultation skills [36]. Our findings confirm that while training is delivered in various ways, its quality and focus are inconsistent across regions. There remains a need to strengthen consultation skills for holistic, person-centred care.

### Implications for research and/or practice

NHS England’s updated SMR guidance for 2025/26 now includes a requirement to ensure patients understand the purpose of SMRs [37]. SMR information resources, such as those developed by the Health Innovation Network [38, 39], include multilingual and easy-read formats and have been reported to help improve uptake in underserved groups. However, persistent engagement gaps remain.

Optimising SMRs means embedding them as routine, holistic reviews that are well-communicated, well-conducted, adequately resourced, and delivered by trained, supported pharmacists. This requires clarity on clinical roles, protected time and consistent pathways. When implemented well, SMRs could have the potential uncover unmet patient needs, support safe deprescribing to reduce medication-related harm, and improve patient understanding of their medicines, ultimately improving outcomes for people with MLTCs.

## Conclusions

This study highlights the key mechanisms shaping the implementation of SMRs into routine practice in primary care. As NHS England continues to refine national guidance, our findings point to both the promise and the pressures of embedding SMRs sustainably. Optimising SMRs will require strategic and sustained investment from policymakers in training, development and support, clearer definitions of clinical pharmacist roles and responsibilities, and national communication strategies to promote awareness and engagement. These steps could help ensure that SMRs are conducted consistently and meaningfully across primary care settings, offering patients not just a transactional review but a personalised and holistic opportunity to optimise their care.

## Data Availability

All data produced in the present study are available upon reasonable request to the authors

## List of abbreviations

ADEs: Adverse drug events
CPPE: Centre for Pharmacy Postgraduate Education
DES: Directed Enhanced Service
ICBs: Integrated Care Boards
IIF: Impact and Investment Fund
GP: General Practitioner
MLTCs: Multiple long-term conditions
NHS: National Health Service
NPT: Normalization Process Theory
NHS: National Health Service
NIHR: National Institute for Health Research
OSCAR: Optimising StruCtured medicAtion Reviews evaluation
PCN: Primary Care Network
SMRs: Structured Medication Reviews

## Ethical Approval

This study received NHS Research Ethics Committee and Health Research Authority approval (reference 22/SC/0373).

## Acknowledgements

We thank Lucy Curtin for administrative support throughout the project, and Dr Adaku Agwunobi for her work in undertaking qualitative interviews with research participants. We are very grateful to all those who dedicated their time to participating in this research. We also wish to thank the members of the OSCAR steering group and Patient and Public Involvement members for their invaluable time and insight provided throughout the project.

## References

1. Parekh, N., Ali, K., Stevenson, J. M., Davies, J. G., Schiff, R., Van der Cammen, T., Harchowal, J., Raftery, J., & Rajkumar, C. (2018). Incidence and cost of medication harm in older adults following hospital discharge: a multicentre prospective study in the UK. Br J Clin Pharmacol, 84(8), 1789–1797. 10.1111/bcp.13613.

2. Elliott, R. A., Camacho, E., Jankovic, D., et al. (2021). Economic analysis of the prevalence and clinical and economic burden of medication error in England. BMJ Qual Saf. 30:96–105.

3. Masnoon, N., Shakib, S., Kalisch-Ellett, L., & Caughey, G. E. (2017). What is polypharmacy? A systematic review of definitions. BMC Geriatrics, 17(1), 230. 10.1186/s12877-017-0621-2.

4. Guthrie, B., Makubate, B., Hernandez-Santiago, V., & Dreischulte, T. (2015). The rising tide of polypharmacy and drug-drug interactions: population database analysis 1995–2010. BMC Medicine, 13(1), 74. 10.1186/s12916-015-0322-7

5. Mangoni, A.A. and Jackson, S.H.D. (2004), Age-related changes in pharmacokinetics and pharmacodynamics: basic principles and practical applications. British Journal of Clinical Pharmacology, 57: 6–14. 10.1046/j.1365-2125.2003.02007.x

6. Doumat, G., Daher, D., Itani, M., Abdouni, L., El Asmar, K., & Assaf, G. (2023). The effect of polypharmacy on healthcare services utilization in older adults with comorbidities: a retrospective cohort study. BMC Primary Care, 24(1), 120. 10.1186/s12875-023-02070-0

7. Davies, L. E., Spiers, G., Kingston, A., Todd, A., Adamson, J., & Hanratty, B. (2020). Adverse Outcomes of Polypharmacy in Older People: Systematic Review of Reviews. Journal of the American Medical Directors Association, 21(2), 181–187. 10.1016/j.jamda.2019.10.022

8. Osanlou, R., Walker, L., Hughes, D. A., Burnside, G., & Pirmohamed, M. (2022). Adverse drug reactions, multimorbidity and polypharmacy: a prospective analysis of 1 month of medical admissions. BMJ Open, 12(7), e055551. 10.1136/bmjopen-2021-055551

9. Zaninotto, P., Huang, Y. T., Di Gessa, G., Abell, J., Lassale, C., & Steptoe, A. (2020). Polypharmacy is a risk factor for hospital admission due to a fall: evidence from the English Longitudinal Study of Ageing. BMC Public Health, 20(1), 1804. 10.1186/s12889-020-09920-x

10. National Institute for Health and Care Excellence. (2015). NICE Guideline 5: Medicines optimisation: the safe and effective use of medicines to enable the best possible outcomes.

11. British Medical Association and NHS England. (2019). Investment and evolution: A five-year framework for GP contract reform to implement The NHS Long Term Plan.

12. NHS England and NHS Improvement. (2020). Network Contract Direct Enhanced Service: Contract specification 2020/21 – PCN Requirements and Entitlements.

13. NHS England. (2020). Network Contract Directed Enhanced Service. Structured medication reviews and medicines optimisation: Guidance 2020-21.

14. NHS England. (2024). Network contract DES guidance for 2024/25: Part A - Clinical and support services (Section 8). https://www.england.nhs.uk/publication/network-contract-directed-enhanced-service-guidance-for-2024-25-in-england-part-a-clinical-and-support-services-section-8/.

15. Duncan, P., Cabral, C., McCahon, D., Guthrie, B., & Ridd, M. J. (2019). Efficiency versus thoroughness in medication review: a qualitative interview study in UK primary care. British Journal of General Practice, 69(680), e190–e198. 10.3399/bjgp19X701321.

16. Stewart, D., Madden, M., Davies, P., Whittlesea, C., McCambridge, J. (2021). Structured medication reviews: origins, implementation, evidence, and prospects. Br J Gen Pract. 71 (709): 340–341. 10.3399/bjgp21X716465.

17. Doherty, A. S., Boland, F., Moriarty, F., Fahey, T., & Wallace, E. (2023). Adverse drug reactions and associated patient characteristics in older community-dwelling adults: a 6-year prospective cohort study. Br J Gen Pract. 73(728), e211–e219. 10.3399/bjgp.2022.0181.

18. Royal Pharmaceutical Society. (2019). Polypharmacy: Getting our medicines right.

19. Royal Pharmaceutical Society. (2013). Medicines optimisation: Helping patients to make the most of medicines.

20. McCahon, D., Denholm, R. E., Huntley, A. L., Dawson, S., Duncan, P., & Payne, R. A. (2021). Development of a model of medication review for use in clinical practice: Bristol medication review model. BMC Medicine, 19(1), 262. 10.1186/s12916-021-02136-9.

21. McEvoy, R., Ballini, L., Maltoni, S., O’Donnell, C. A., Mair, F. S., MacFarlane A. (2014). A qualitative systematic review of studies using the normalization process theory to research implementation processes. Implement Sci. 2;9:2. doi: 10.1186/1748-5908-9-2.

22. Agwunobi et al., (under review). Understanding Structured Medication Reviews delivered by primary care pharmacists in England: a national cross-sectional survey. BMJ Open.

23. Saunders, B., Sim, J., Kingstone, T., Baker, S., Waterfield, J., Bartlam, B., Burroughs, H., Jinks, C. (2018). Saturation in qualitative research: exploring its conceptualization and operationalization. Qual Quant. 52(4):1893–1907. doi: 10.1007/s11135-017-0574-8.

24. May, C.R., Albers, B., Bracher, M., Finch, T., Gilbert, A., Girling, M., Greenwood, T., MacFarlane, A., Mair, F. S., May, C. M., Murray, E., Potthoff, S., Rapley, T. (2022). Translational framework for implementation evaluation and research: a normalisation process theory coding manual for qualitative research and instrument development. Implementation Sci 17, 19. 10.1186/s13012-022-01191-x.

25. May, C. R., Cummings, A., Girling, M., Bracher, M., Mair, F. S., May, C. M., Murray, E., Myall, M., Rapley, T., Finch, T. (2018). Using Normalization Process Theory in feasibility studies and process evaluations of complex healthcare interventions: a systematic review. Implementation Sci 13, 80. 10.1186/s13012-018-0758-1.

26. Braun, V., & and Clarke, V. (2006). Using thematic analysis in psychology. Qualitative Research in Psychology, 3(2), 77–101. 10.1191/1478088706qp063oa.

27. Campbell, R., Goodman-Williams, R., Feeney, H., & Fehler-Cabral, G. (2020). Assessing Triangulation Across Methodologies, Methods, and Stakeholder Groups: The Joys, Woes, and Politics of Interpreting Convergent and Divergent Data. American Journal of Evaluation, 41(1), 125–144. 10.1177/1098214018804195.

28. General Pharmaceutical Council. (2023). The GPhC register as of 30 September 2023 – Diversity data tables – England.

29. Baird, B., Lamming, L., Bhatt, R. T., Beech, J., Dale, V. (2022). Integrating additional roles into primary care networks. The King’s Fund: London.

30. Turk, A., Wong, G., Mahtani, K. R., Maden, M., Hill, R., Ranson, E., Wallace, E., Krska, J., Mangin, D., Byng, R., Lasserson, D., & Reeve, J. (2022). Optimising a person-centred approach to stopping medicines in older people with multimorbidity and polypharmacy using the DExTruS framework: a realist review. BMC Medicine, 20(1), 297. 10.1186/s12916-022-02475-1.

31. Mann, C., Anderson, A., & Boyd, M. (2022). The role of clinical pharmacists in general practice in England: Impact, perspectives, barriers and facilitators. Research in Social and Administrative Pharmacy, 18(8), 3432–3437. 10.1016/j.sapharm.2021.10.006.

32. Anderson, C., Zhan, K., Boyd, M., Mann, C. (2019). The role of pharmacists in general practice: a realist review. Res Social Adm Pharm. 15: 338–45.

33. Bradley, F., Seston, E., Mannall, C., Cutts, C. (2018). Evolution of the general practice pharmacist’s role in England: a longitudinal study. Br J Gen Pract, DOI: 10.3399/bjgp18X698849.

34. Madden, M., Mills, T., Atkin, K., Stewart, D., McCambridge, J. (2022). Early implementation of the structured medication review in England: a qualitative study. Br J Gen Pract. 20;72(722):e641–8. doi: 10.3399/BJGP.2022.0014.

35. Swinglehurst, D., Hogger, L., Fudge, N. (2023). Negotiating the polypharmacy paradox: a video-reflexive ethnography study of polypharmacy and its practices in primary care. BMJ Quality & Safety. 32:150–159.

36. Madden, M., Stewart, D., Mills, T., McCambridge, J. (2023). Consultation skills development in general practice: findings from a qualitative study of newly recruited and more experienced clinical pharmacists during the COVID-19 pandemic. BMJ Open. 13:e069017. doi: 10.1136/bmjopen-2022-069017.

37. NHS England. (2025). Network contract directed enhanced service – guidance for 2025/26 in England – part A: clinical and support services (section 8). https://www.england.nhs.uk/publication/network-contract-directed-enhanced-service-guidance-for-2025-26-in-england-part-a-clinical-and-support-services-section-8/.

38. Health Innovation Network. (2023). Resources to support patients having a Structured Medication Review. https://thehealthinnovationnetwork.co.uk/programmes/medicines/polypharmac y/patient-information/.

39. Health Innovation Network. (2025). Improving access to Structured Medication Reviews in seldom-heard communities: Understanding the enablers and barriers to addressing health inequalities.

