## Supplementary document A for "Exploring the implementation and integration of structured medication reviews in primary care: A qualitative evaluation using normalization process theory"

Optimising StruCtured medicAtion Review (OSCAR) Qualitative study

**HEALTHCARE PROFESSIONALS INTERVIEW TOPIC GUIDE**

*The interviews will be responsive to participants’ own agendas but will cover the broad*

*topics/questions below. It is common in qualitative work to iteratively develop topics and questions as new ideas emerge from early data collection. Therefore, we may add new topics as the interviews progress and data collection continues. Interviewees will include pharmacists, GP’s, GP practice staff, and care home staff.*

Part 1: Introduction

- Re-introduce self and purpose of call/visit;
- Check with participant:
  - that they are still willing to be interviewed and for the interview to be recorded;
  - duration of interview (30-60 minutes)
  - they can halt the interview at any point
- Check demographic information.

Part 2: Interview

- **Explore healthcare professional experiences of implementing SMR’s (how are they conducted i.e. remotely, in person etc.)**
- **Explore how/why healthcare professionals prioritise patient groups for an SMR**
- **Explore how healthcare professionals prepare for an SMR, and how they prepare patients for an SMR**
- **Explore barriers to conducting SMR’s**
- **Explore healthcare professional’s thoughts on the impact of SMR’s on the population they are targeting**

Part 3: Closing

- Thank participant for taking part in the interview.
- Revisit consent
- Ask if participant has any questions.
- Let them know that you will be sending all participants a summary of study findings.
- Thank participant again for taking part in the interview.
