## Supplementary document B for "Exploring the implementation and integration of structured medication reviews in primary care: A qualitative evaluation using normalization process theory"

Optimising StruCtured medicAtion Review (OSCAR) Qualitative study

**LEADERS/MANAGERS INTERVIEW TOPIC GUIDE**

*The interviews will be responsive to participants’ own agendas but will cover the broad*

*topics/questions below. It is common in qualitative work to iteratively develop topics and questions as new ideas emerge from early data collection. Therefore, we may add new topics as the interviews progress and data collection continues. Interviewees will include leaders and managers from Primary Care Networks (PCN’s) who have been conducting structured medication reviews. The aim of these interviews is for leaders and managers to reflect on the different kinds of work required to implement structured medication reviews at the PCN and individual practice level.*

Part 1: Introduction

- Re-introduce self and purpose of call/visit;
- Check with participant:
  - that they are still willing to be interviewed and for the interview to be recorded;
  - duration of interview (30-60 minutes)
  - they can halt the interview at any point
- Check demographic information.

Part 2: Interview

- Discuss how the roll out of SMR’s have gone so far and the impact of the COVID-19 pandemic
- Discuss issues with the implementation of PCN’s and/or the national network DES specification
- Explore the availability of pharmacists to undertake SMR’s, and whether other healthcare professionals may be tasked with them
- Explore prescribing certification for the pharmacists

Part 3: Closing

- Thank participant for taking part in the interview.
- Revisit consent
- Ask if participant has any questions.
- Let them know that you will be sending all participants a summary of study findings.
- Thank participant again for taking part in the interview.
